# Development of a digital twin for the diagnosis of cardiac perfusion defects

**DOI:** 10.1101/2025.02.04.25321638

**Authors:** Elisabetta Criseo, Andrea Baggiano, Giovanni Montino Pelagi, Guido Nannini, Alberto Redaelli, Gianluca Pontone, Christian Vergara

**Affiliations:** Cardio Tech-lab, Centro Cardiologico Monzino, Via Privata Carlo Parea 4, Milano, 20138, Italy; Dipartimento di Elettronica Informazione e Bioingeneria, Politecnico di Milano, Via Ponzio 34/5, Milano, 20133, Italy; Dipartimento di Cardiologia Peri-operatoria e Imaging Cardiovascolare, Centro Cardiologico Monzino, Via Privata Carlo Parea 4, Milano, 20138, Italy; LaBS, Dipartimento di Chimica, Materiali e Ingegneria Chimica ”Giulio Natta”, Politecnico di Milano, Piazza Leonardo da Vinci, 32, Milano, 20133, Italy; Department of Biomedical, Surgical and Dental Sciences, University of Milan, Milano, Italy

**Keywords:** digital twin, myocardial blood flow, coronary artery disease, computational fluid dynamics

## Abstract

Myocardial Blood Flow (MBF) is a key indicator of myocardial perfusion, typically assessed through additional clinical tests like dynamic CT perfusion under stress. This study introduces a digital twin designed to enhance coronary artery disease diagnosis by predicting MBF using data from routine CT images and clinical measurements. The digital twin employs AI methods to reconstruct coronary and myocardial geometries and integrates a computational model, featuring 3D coronary arteries and a three-compartment myocardial model, blindly calibrated with data from six representative patients. Validation on 28 additional patients showed MBF predictions consistent with experimental and clinical measurements. Confusion matrix analysis assessed the twin’s ability to classify at-risk patients (averaged MBF ***<*** 230 ml/min/100g) versus non-at-risk patients, yielding a recall of 0.77, with precision and accuracy at 0.72. This work represents the first attempt to predict and validate MBF on such a large cohort, paving the way for future clinical applications.

## 1 Introduction

Myocardial perfusion is the process which delivers nutrients and oxygen to the heart muscle, i.e. the myocardium, by means of blood flow in the coronary arteries and microvasculature. This mechanism could be impaired by the stenosis of the coronary arteries lumen, a pathological condition known as Coronary Artery Disease (CAD), which can lead, depending on the stenosis gravity, to *ischemia* or even infarction [1].

The use of only anatomical information, retrieved, for example, by Coronary Computed Tomography Angiography (CCTA), provides only partial insight on CAD, as it is not able to assess the presence of myocardial ischemia [2]. For this reason, the addition of functional information, such as Fractional Flow Reserve (FFR), has been extensively investigated [3, 4]. However, FFR provides information at the large coronary level, proximally to the myocardial region of interest, regarding the functional relevance of epicardial disease, thus not accounting for microvascular disease.

Clinical tests that provide hemodynamic information directly at the myocardial level have recently been introduced; these provide informations regarding both epicardial and microvascular disease. In this regard, stress Computed Tomography Perfusion (stress-CTP) [5–7] is an imaging technique that allows the retrieval of functional information also from the microvasculature. The addition of stress-CTP to the CCTA examination has led to an improvement in the diagnostic protocol for CAD, specifically allowing a more accurate identification of functionally relevant stenosis [8] and yielding an improvement of the positive predictive power [9]. Nevertheless, stress-CTP still presents some drawbacks as patients undergo pharmacologically induced stress conditions and additional radiation exposure after routinary CT [7].

In the last decade, the use of *digital twins* (personalized computational models) has been introduced and investigated to improve the evaluation of stenoses functional relevance and to be an alternative to invasive exams, such as FFR. An effective example of this approach is FFR_CT_, a computational method that allows computing FFR from non-invasive imaging by exploiting Computational Fluid Dynamics (CFD) [10–14].

In recent years, the idea of analyzing also myocardial perfusion, i.e. Myocardial Blood Flow (MBF), through computational models has been gaining interest. In particular, in Table 1 we sum up, to the best of our knowledge, all computational studies that, starting from clinical data and using patient-specific data, aimed to predict MBF and assess its precision with respect to clinical MBF measures. Most of them use the assumption of a fluid in a porous medium to describe the hemodynamics in the myocardium [15], by means of either a single-compartment [16–18] or a multi-compartment Darcy model [19–23]. In one paper by our group [21] a surrogate model of compliance has been proposed to recover the typical diastolic nature of the coronary flow rate. In all the cases analysed the myocardial Darcy model is coupled with either a 1D or 3D model of the coronary arteries, allowing to include proximal inputs and geometrical information on coronary stenosis.

**Table 1:** State of the art of computational studies aiming at reproducing or predicting MBF with application to clinical data; we distinguish between blindly and non-blindly calibrated models. NS = Navier-Stokes, FSI = Fluid Structure Interaction, 1C = one Compartment, 3C = three Compartment, rDarcy = 3D rigid Darcy, cDarcy = 3D compliant Darcy, LPM = Lumped Parameters Model, PS = Patient Specific, FR = Flow Rate, AoP = Aortic Pressure, *Volume Based* = Calibration based on geometrical volume information, *Region Based* ✓ = Specific calibration for each perfusion region, *Region Based ×* = Uniform calibration (constant parameters in space), *Groups* = Number of patients’ groups (based on the degree of stenosis) for which a specific calibration is performed, MBF = Myocardial Blood Flow, oCAD = obstructive CAD.

|  |  | Not blinded calibration |  | Blinded calibration |  |  |  |  |
| --- | --- | --- | --- | --- | --- | --- | --- | --- |
|  |  | DiGregorio (2021, 2022) [20, 23] | Menon (2024) [18] | Papamanolis (2021) [16] | Kim (2023) [17] | Montino P. (2024) [22] | Montino P. (2024) [21] | Present work |
| **Mathematical Model** | Coronaries | 3D NS | 3D FSI | 1D FSI | 1D FSI | 3D NS | 3D NS | 3D NS |
|  | Microvasculature | rDarcy-3C | rDarcy-1C + closed-loop LPM | rDarcy-1C + LPM | rDarcy-1C + windkessel | rDarcy-3C | cDarcy-3C | cDarcy-3C |
| **Inlet BCs** |  | PS FR | closed-loop LPM | PS FR | PS FR | PS AoP | PS AoP | PS AoP |
| **Calibration** | Volume Based | ✓ | ✓ | ✓ | ✓ | × | × | ✓ |
|  | Patient-Specific | ✓ | ✓ | ✓ | × | × | × | ✓ |
|  | Region Based | ✓ | × | × | × | × | × | ✓ |
|  | Groups | 1 | 1 | 1 | 1 | 1 | 1 | 2 |
| **Output** |  | MBF map | MBF map | MBF bullseye view | MBF map | in-space-average MBF | MBF map | MBF map |
| **#Patients** |  | 8 | 6 | 5 | 1 | 7 | 1 | 28 |
| **Type** |  | oCAD & non-oCAD | oCAD | non-oCAD | non-oCAD | oCAD & non-oCAD | non-oCAD | oCAD & non-oCAD |

One of the most challenging issues related to the cardiac perfusion Darcy model is the calibration of its parameters (conductances and permeabilities), which are highly patient dependent and difficult to measure. Noticeably, some of the cited works took significant steps towards a blind calibration of such parameters, i.e. without using *a priori* any MBF clinical measure [16, 17, 21, 22].

In this paper, we propose a new digital twin resulting from the combined use of Artificial Intelligence (AI) and a computational model [21, 22], for the reconstruction of coronary arteries and myocardium geometries and for the prediction of myocardial perfusion, respectively (see Fig. 1). Specifically:

**Fig. 1:**
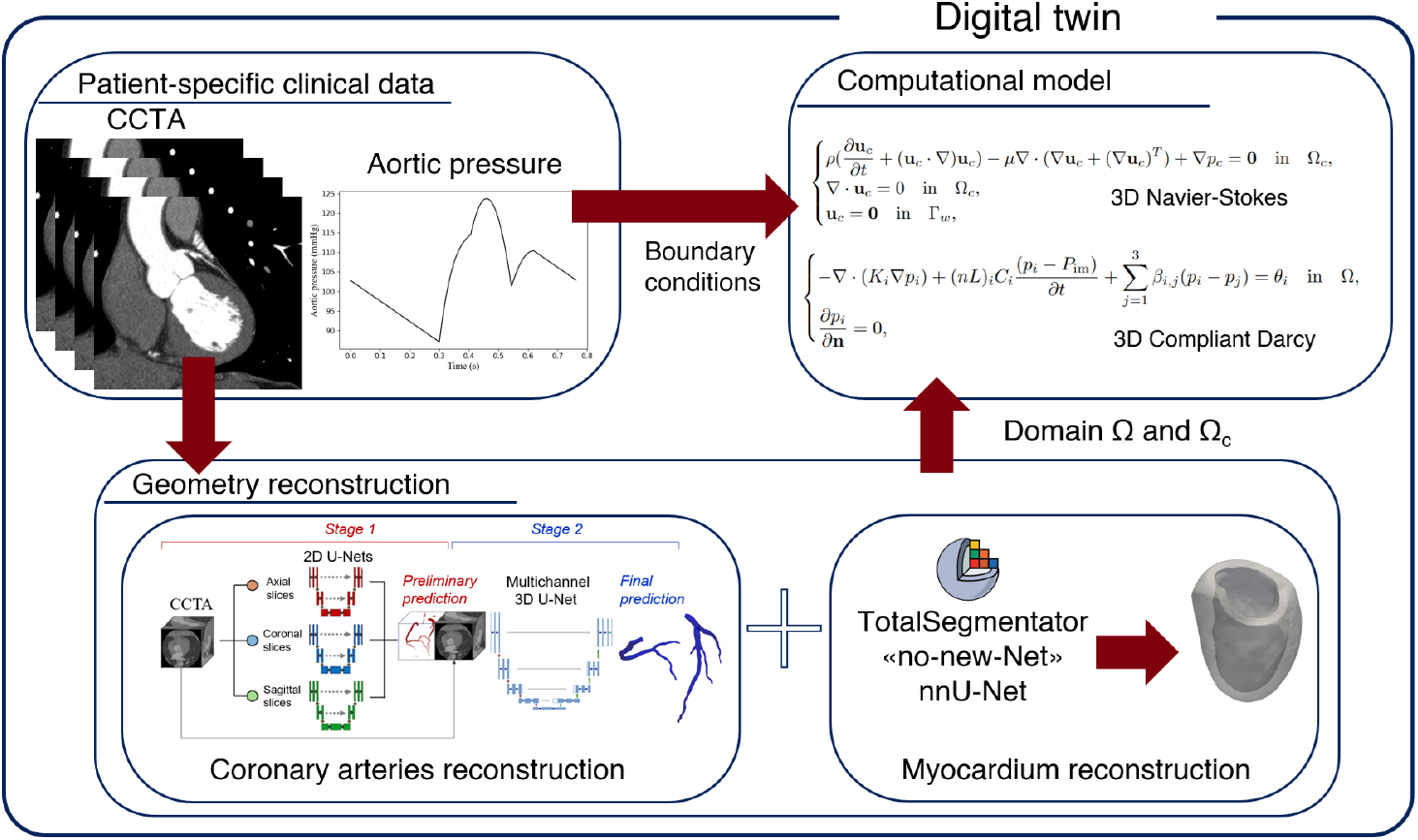
Graphical representation of the proposed digital twin which features the use of patient-specific data both to reconstruct the coronaries and myocardium and to impose boundary conditions to the computational model for MBF prediction. For geometry reconstruction both in house [24] and open-source AI (U-Net) models are used [25].

- We perform the segmentation of the coronary arteries and myocardium geometries from the patients’ CCTA images in less than 15 minutes;
- We use a computational model (developed by the research group in [21]) based on the coupling between 3D incompressible fluid dynamics for the large coronaries and a three-compartment compliant Darcy model for the myocardial microcirculation;
- We calibrate such computational model with a region and volume based approach using anatomical data from six representative patients; then we apply the calibration to the remaining 28 patients, which are used to test the digital twin accuracy.

Through these steps, our aim is to provide an accurate prediction of MBF indicators that describe myocardial perfusion and to validate our results against the patients’ data.

This work introduces, to our knowledge, the first digital twin for MBF prediction which is blindly calibrated and applied to a wide cohort of patients.

## 2 Methods

In this section we want to provide details about the clinical dataset used for this work (Section 2.1). Moreover, we describe the approach used for geometry reconstruction in Section 2.2, the boundary conditions formulation and setting of numerical simulations in Section 2.3. In Section 2.4, we describe the process of the digital twin parameters calibration and, finally, in Section 2.5 we describe the process used for the results analysis.

### 2.1 Clinical dataset

We retrieved both CCTA and stress-CTP scans of 34 patients from the PERFECTION study dataset [6, 26]. Rest CCTA images were performed using a Revolution CT scanner (GE Healthcare, Milwaukee, Wisconsin) according to the recommendations of the Society of Cardiovascular Computed Tomography (SCCT) [27]. Stress-CTP images were acquired after inducing a pharmacological stress by the means of adenosine injection. Additional information on CCTA and stress-CTP acquisition protocols can be found in [6]. The patients selected had symptoms of myocardial ischemia (such as *angina pectoris*) but no prior history of myocardial infarction, revascularization procedure of the coronary arteries or acute coronary syndrome; the main characteristics of the cohort used in the present work are summed up in Table 2.

**Table 2:** Main characteristics of the cohort of patients selected for this work. The values are expressed in ”mean *±* standard deviation” or ”number (%)”.

| Baseline characteristics |  | Risk factors |  | Clinical measurements |  |
| --- | --- | --- | --- | --- | --- |
| Number of patients | 34 | Family history | 22 (65%) | Systolic pressure | 144 ± 15 |
| Age (years) | 65 ± 8 | Smoking | 14 (41 %) | Diastolic pressure | 81 ± 10 |
| Men | 27 (79%) | Hypertension | 24 (71%) | Heart Rate | 57 ± 9 |
| Height | 171 ± 8 | Dyslipidemia | 22 (65%) | | |
| Weight | 77 ± 15 | Diabetes | 6 (18%) | | |
| Body Mass Index (kg/m<sup>2</sup>) | 27 ± 5 | | | | |

### 2.2 Geometry reconstruction

Rest CCTA scans are used to reconstruct both the patients’ myocardium and coronary arteries. In what follows we provide some details about the geometry reconstruction and the generation of the perfusion regions.

#### 2.2.1 Coronary arteries reconstruction

Our starting point is the raw coronary arteries segmentations. The lumen of the vessel was reconstructed from CCTA images, by means of a neural network based on the the nn-UNet framework [28], described in [24], developed at DEIB, Politecnico di Milano. The network was trained to segment the three main coronary branches (i.e., the Left Anterior Descending (LAD), the Left CircumfleX (LCX) and the Right Coronary Artery (RCA)) and include sub-branches with caliber larger than 1.5 mm. The resulting dataset consists of geometries that are characterized by about seven/eight outlets corresponding to the main coronary branches.

These are then processed, using VMTK [29] (http://www.vmtk.org/) and Paraview (https://www.paraview.org/), to improve their geometric details in view of the computational meshes. In particular, we perform the following steps (see also Fig. 2):

**Fig. 2:**
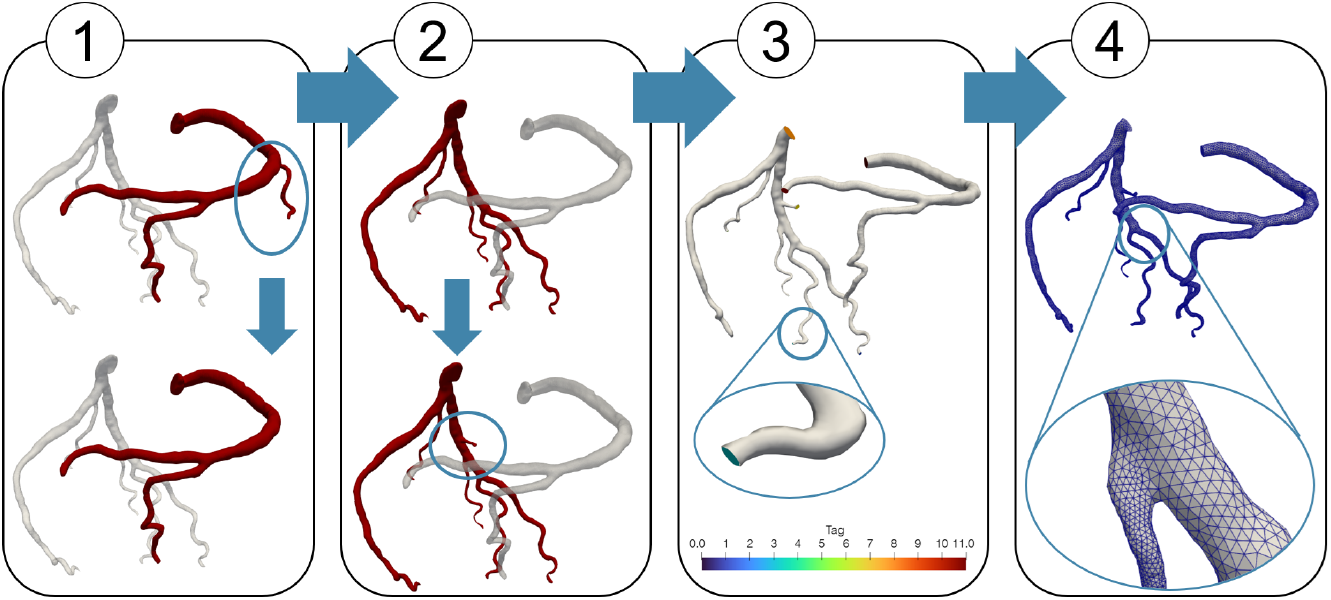
Main steps for coronary geometry reconstruction after segmentation with the cascaded CNN: 1) remove the right coronary branches that perfuse the right ventricle; 2) add new branches if necessary; 3) tag each coronary inlet and outlet; 4) create the non-uniform computational mesh.

1. we remove the right coronary branches which perfuse the right ventricle, as we do not include it in the final computational domain;
2. we compare the reconstructed geometry with what we observe from the CCTA and, if necessary, we segment some additional branches;
3. we smooth the geometry and assign to each inlet and outlet a specific tag;
4. we create non-uniform tetrahedric computational meshes, where the elements have different dimensions depending on the radius of the vessel.

The computational time to reconstruct each geometry and build the corresponding computational mesh is about 30 minutes.

#### 2.2.2 Myocardium reconstruction

As for the reconstruction of the patients’ myocardium geometries, we perform an automatic segmentation exploiting the TotalSegmentator tool [25] on 3D Slicer (https://www.slicer.org/). We consider only the left ventricle identified as that region below the valves plane, which represents the ventricle base. The segmented geometry is then processed using VMTK, specifically we assign different tags to the left ventricle endocardium, epicardium and base; then we mesh the geometry using tetrahedric elements of constant length h=1.5mm.

#### 2.2.3 Perfusion regions generation

As a reliable assumption, we can consider that each reconstructed coronary branch feeds only one specific myocardial region and, vice versa, that each myocardial region is fed by only one large coronary [30]. Accordingly, the computational model considered in this work, see Section 2.3, is based on identifying the *perfusion regions*, whose union covers the entire myocardium and each of which is associated with a specific large coronary outlet.

The personalized perfusion regions are generated starting from the patient-specific coronary arteries geometry. In particular, we solve an *Eikonal problem* [20] that has the aim of identifying in the myocardium the closest points to a specific coronary outlet, which is distinguished based on the tag assigned (Fig. 3). This process allows to evaluate the volume of region perfused by a reconstructed coronary branch and to assign different parameters to each region. The time taken for myocardial reconstruction and generation of perfusion regions is about 1 hour.

**Fig. 3:**
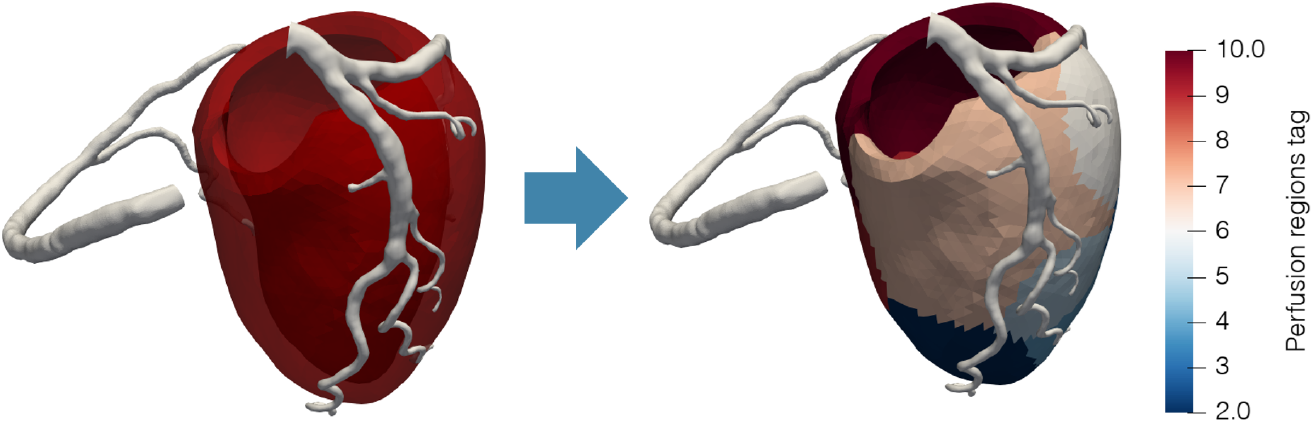
Generation of the perfusion regions based on the correspondence with the coronary outlets.

### 2.3 Building the digital twin: Mathematical modeling and numerical setting

For the construction of the digital twin and the computation of the myocardial blood flow maps, we use the mathematical and computational model proposed in [21]. This is based on a three-compartment Darcy model for intramyocardial hemodynamics, representing the small coronary arteries (compartment I), arterioles (compartment II) and capillaries (compartment III), coupled with incompressible Navier-Stokes equations for the large coronaries. The Darcy problem has been modified to account for variation in the dimensions of pores and thus for systolic contraction.

Our digital twin requires geometric and functional data as an input. The former are provided by the reconstructed geometries and perfusion regions described above, whereas the latter consist in pressure inlet conditions at the coronary ostia. Such boundary conditions are based on the prescription of a personalized aortic pressure as described in [22] (Fig. 4, left).

**Fig. 4:**
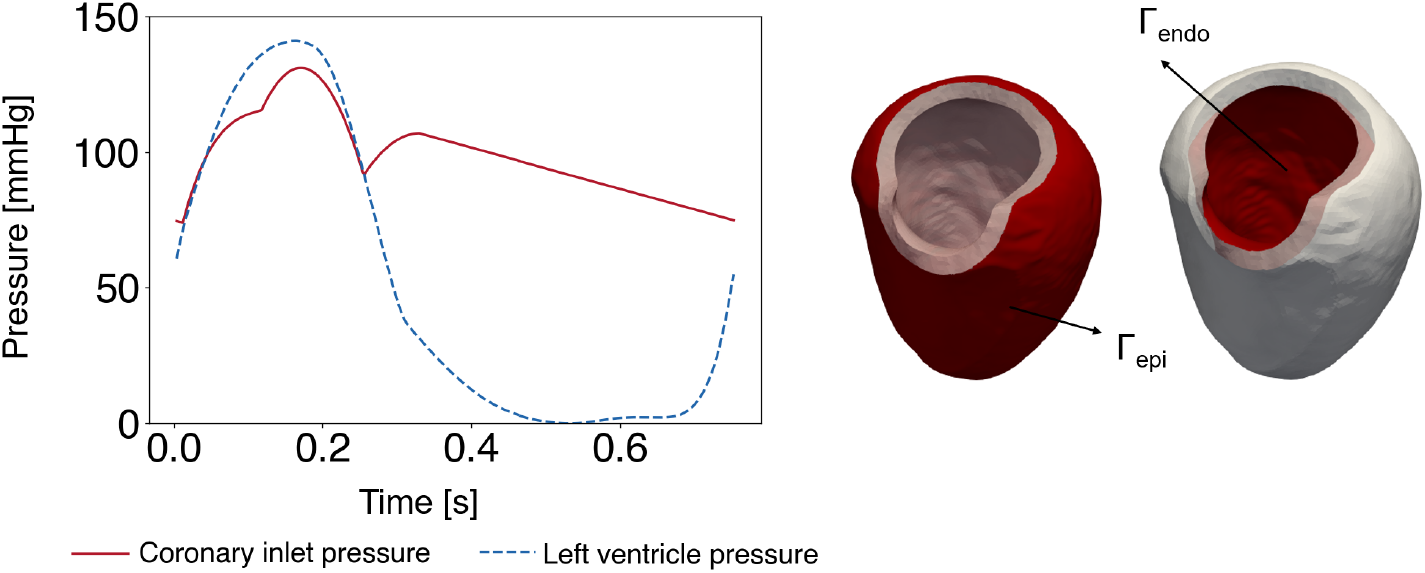
Left: an example of the personalized pressure curve imposed as boundary condition to the coronary inlets (in red) and the approximation of the left ventricle pressure used to obtain P_IM_ (in blue). Right: representation of Γ_*epi*_ and Γ_*endo*_, the epicardial and endocardial wall respectively.

In order to account for the vessel distensibility the parameter *C*_*i*_ was introduced, in [21], and defined as follows for the i-th compartment:

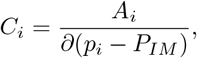

where *A*_*i*_ is the average cross sectional area of vessels in the i-th compartment. The vessel distensibility is a time-dependent parameter and, in particular, is influenced by the time-variation of the intramyocardial pressure *P*_*IM*_, which varies linearly along the transmural direction (from the epicardium to the endocarium surface) and is defined as follows:

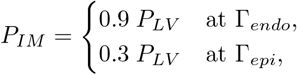

where Γ_*endo*_ and Γ_*epi*_ are the endocardial and the epicardial wall, respectively (Fig. 4, right). Moreover, *P*_*LV*_ is the left ventricle pressure, which has been obtained through a preliminary electromechanics simulation performed as in [31] and personalized for each patient as in [22] (see Fig. 4, left).

For each patient, we compute the 3D blood velocity and pressure both for the coronaries and the myocardium during four cardiac cycles, with a time step Δ*t* = 2 *·* 10^*−*3^ s and a personalized period based on the patient’s data. Then we discard the first three cardiac cycles and we analyze only the last one, as we deem periodicity to be reached.

We solve the numerical discretization of the mathematical problem using life^x^ [32] (https://lifex.gitlab.io/), a high-performance library for the finite element simulations of multi-physics, multiscale, and multi-domain problems, developed within the iHeart project (https://iheart.polimi.it/), at the MOX laboratory of the Department of Mathematics — Politecnico di Milano. Digital twin simulations are run in parallel on 56 cores with CPUs Xeon Gold 6238R@2.20GHz and the computational time requested for simulation of four cardiac cycles is about 1.5 hours.

### 2.4 Building the digital twin: Calibration of the model

The digital twin is governed by 16 functional parameters. Eight of them could be measured or directly obtained from the literature. Specifically, we refer to the quantities listed in Table 3. The length density (nL)_*i*_ is the density of vessels belonging to compartment i (i=1,2,3) that have an average length L, where n is the local density of vessels in the i-th compartment (ratio between the number of vessels in the compartment and compartment volume). The specific permeability *κ* appears in the computation of compartment permeabilities *K*_*i*_ = (*κ*_*i*_*/µ*)(*nL*)_*i*_*A*_*i*_, where *A*_*i*_ is the average cross sectional area of vessels in the i-th compartment.

**Table 3:** Parameters used for the digital twin simulations; quantities that change over the three compartments are indicated with the subscript *i* and their values are reported for each compartment.

| Parameters | Value | Reference |
| --- | --- | --- |
| **Length density $(nL)_i$** [mm mm<sup>-3</sup>] | $nL_1 = 5 \cdot 10^{-1}$; $nL_2 = 1.5 \cdot 10^1$; $nL_3 = 8 \cdot 10^3$ | [33–35] |
| **Specific permeability $\kappa_i$** [m<sup>2</sup>s<sup>-1</sup>Pa<sup>-1</sup>] | $\kappa_1 = 5 \cdot 10^{-9}$; $\kappa_2 = 5 \cdot 10^{-9}$; $\kappa_3 = 5 \cdot 10^{-9}$ | [19] |
| **Blood density $\rho$** [kg m<sup>-3</sup>] | $1.06 \cdot 10^3$ | |
| **Blood viscosity $\mu$** [Pa s] | $3.5 \cdot 10^{-3}$ | |

The other eight parameters of the digital twin were neither available in literature nor measurable. For this reason, they were obtained through an iterative calibration process. Since the purpose of the digital twin is, eventually, using it as a diagnostic tool, our calibration needs to be blind, i.e. it should not directly exploit the MBF maps measured on the patients.

To create an accurate and versatile digital twin, we identify distinct patient groups, based on anatomical features, and we perform a tailored calibration on one representative patient for each group, referred in what follows to as *calibration patient*. The hypothesis is that our calibration works well also for all those (non-calibrated) patients with similar characteristics, referred in what follows to as *validation patients*.

We first divide the cohort at our disposal based on the degree of stenosis of the LAD, LCX and RCA, described on the medical report and observable from the CCTA:

- Group A: patients where the stenosis degree in LAD, LCX and RCA is less than 70% and where the clinicians have not identified an induced ischemia;
- Group B: patients where the stenosis degree in at least one among LAD, LCX and RCA is more than 70% or where the clinicians have identified an induced ischemia,

where for stenosis degree we mean the normalized difference between the diameter of the stenosis D_S_ and the upstream diameter 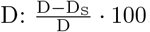.

Secondly, we identify six groups, each with specific characteristics in terms of perfusion regions volume, distinguished by different values of the following volume ratios:

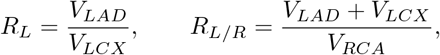

where by *V*_*X*_ we mean the volume perfused by the vessel *X*. These ratios provide insight into how the myocardium is split into the generated perfusion regions and specifically into the relative size of the myocardium perfused by each main coronary. This information results from defining the perfusion regions using an Eikonal problem [20] which incorporates information about the coronary radii.

The definitions of the six groups are reported in Fig. 5, top. The cut-off values for *R*_*L*_ and *R*_*L/R*_ were chosen analyzing their values for all 34 patients (reported in Fig. 5, bottom) and aiming an homogeneous distribution of the patients among the different groups. The groups B-I and B-II are defined according only to the value of *R*_*L/R*_ since, for group B, most of the values of *R*_*L*_ are very similar, apart from two outliers (*R*_*L*_ = 11.2 and 15.3).

**Fig. 5:**
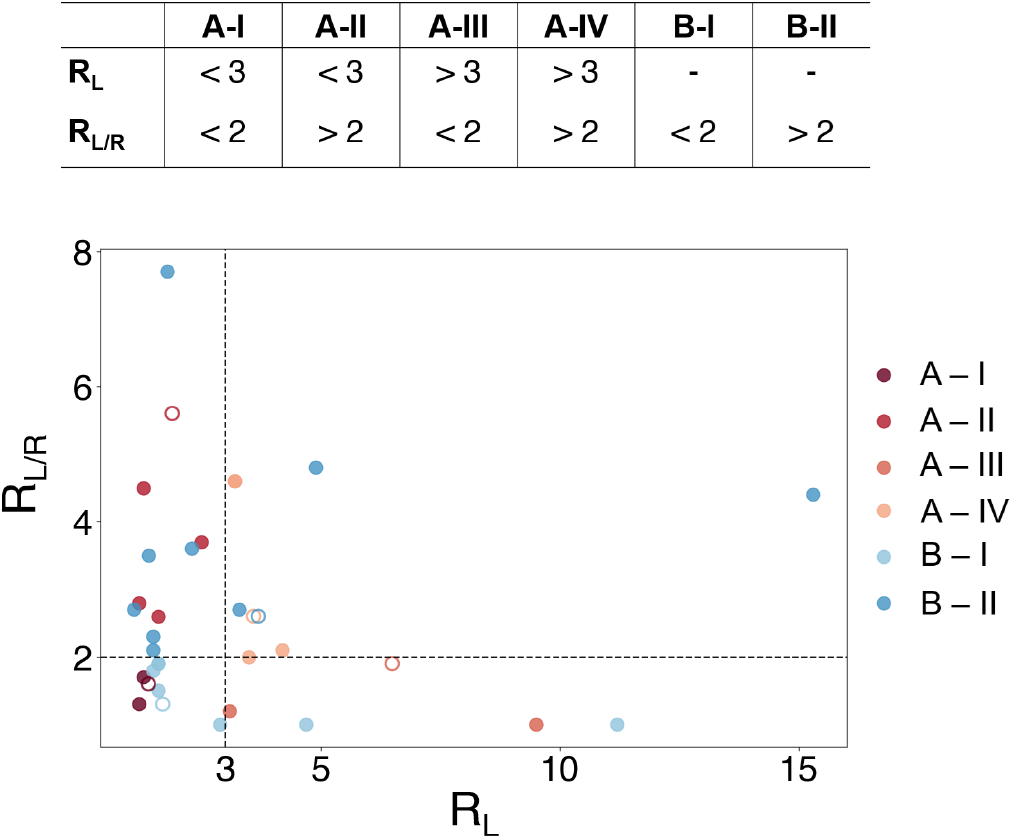
Definition of the six calibration groups (top) and graphical representation of the ratios *R*_*L*_ and *R*_*L/R*_ for each patient (bottom), the dotted lines represent the cut-off used to identify the six groups; hollow dots represent the *calibration patients*.

As a consequence, we perform six different calibrations of the model, one for each group. The calibrated parameters, all constant in time and defined in [21], are the following:

- The morphometry factor *δ*_*i*_, which has a different value for each compartment *i* = 1, 2, 3, influencing the exchange of blood among the three compartments;
- The coupling coefficient *α*, that regulates the exchange of blood from the coronaries to the myocardium; we differentiate its value according to the coronary outlet, i.e. we introduce three different values of *α*, corresponding to LAD, LCX, RCA;
- The constant veins conductance *γ*, part of the sink term, which describes the venous return;
- The constant right atrium pressure *p*_*ra*_, adjusted coherently with *γ* to describe the venous return.

For the calibration of such parameters we introduce, for each calibration patient, the following quantities, where *j* = 1, …, *N* identifies a specific perfusion region:

- *S*_*j*_ the in-space-averaged MBF obtained with the digital twin simulation;
- *M*_*j*_ the in-space-averaged MBF obtained from the stress-CTP;
- 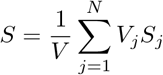 and 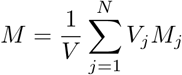 the average of *S* _*j*_ and *M* _*j*_ weighted with the perfusion regions volume *V*_*j*_, where *V* the total myocardial volume;
- 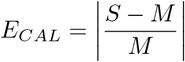 the global relative calibration error.

As for the calibration of all parameters (*δ*_*i*_, *γ* and *p*_*ra*_ and the three *α*) we update iteratively their values aiming at the reduction of the calibration error *E*_*CAL*_ under the value of 5% for the non-severe stenotic patients (group A) and of 10% for group B due to their increased complexity. We used a basic direct search method based on the Hooke and Jeeves algorithm [36] and on an update at each iteration based on the computed *E*_*CAL*_.

### 2.5 Post-processing of results

Once the model is calibrated in accordance with what described in the previous section, we identify for each *validation patient* their corresponding group, according to the subdivision reported in Fig. 5. Then, we apply our calibrated digital twin to each of the *validation patients*, and we post-process the results in view of their comparison with stress-CTP maps. To this aim, we consider average quantities performed over the perfusion regions. Specifically, we compute *S*_*j*_ and *M*_*j*_ and, accordingly, *S* and *M*, as described in Section 2.4. Finally, we define the *validation error E*_*V AL*_ as a measure of the error between the digital twin prediction and the stress-CTP map:

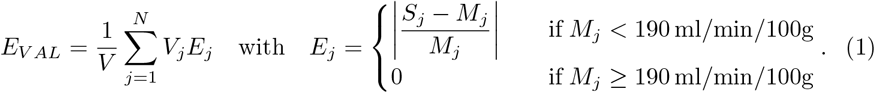

Notice that in the definition of *E*_*j*_ we introduce a cut-off, such that for regions where MBF is greater than 190 ml/min/100g we set the corresponding error *E*_*j*_ to 0, since, for clinical purposes, the accuracy in such regions is not relevant.

### 2.6 Statistical analysis

Statistical analysis is carried out using the Python SciPy 1.14.1 statistics library [37]. Suitable tests to check normal distribution of the values are first performed, specifically Kolmogorov-Smirnov tests. Then we move to the analysis of the correlation between the the stress-CTP measures and digital twin results; to this aim we perform Pearson test for correlation between *S* and *M* (for which the number of samples is small) and Spearman test for correlation between *S*_*j*_ and *M*_*j*_ (for which the number of samples is larger).

The agreement between the digital twin results and clinical measures is evaluated using Bland-Altman analysis, while diagnostic accuracy of the digital twin is assessed using *confusion matrix analysis*. This is based on the definition of a threshold *η* for the evaluation of the model performance considering four categories: true positive (TP, *M < η* & *S < η*), true negative (TN, *M > η* & *S > η*), false positive (FP, *M > η* & *S < η*) and false negative (FN, *M < η* & *S > η*). As indicator of performance we use: the true negative rate (or specificity) TNR = #*TN/*(#*TN* + #*FP*), which expresses the performance of the model to classify samples over the established threshold; the true positive rate (or sensitivity) TPR = #*TP/*(#*TP* + #*FN*), which expresses how many of the relevant values are retrieved; the precision = (#*TP/*(#*TP* + #*FP*), which is the ability of the model to identify how many of the retrieved values are relevant; the accuracy = (#*TP* + #*TN*)*/*(#*FP* + #*FN* + #*TP* + #*TN*), which expresses the ability of the method to well classify the patients in the correct category.

For all tests, a *p <* 0.05 was considered statistically significant.

## 3 Results

### 3.1 Model calibration

We start by reporting the results of the calibration of our computational model. In Table 4 we list all the calibrated parameters and we report the calibration error *E*_*CAL*_ obtained for each *calibration patient*. For all the groups (except for B-I), the obtained *E*_*CAL*_ is much smaller than the threshold used as stopping criterion during the calibration procedure (see Section 2.4).

**Table 4:** Calibrated parameters for each group and calibration error *E*_*CAL*_ after running the digital twin simulations with the calibrated parameters for each group.

|  | A - I | A - II | A - III | A - IV | B - I | B - II |
| --- | --- | --- | --- | --- | --- | --- |
| **Morphometry factors** $\delta_i$<br>[mm<sup>-1</sup> s<sup>-1</sup> Pa<sup>-1</sup>] | $\delta_1 = 5 \cdot 10^{-3}$<br>$\delta_2 = 5 \cdot 10^{-2}$<br>$\delta_3 = 1 \cdot 10^1$ | $5 \cdot 10^{-3}$<br>$5 \cdot 10^{-2}$<br>$1 \cdot 10^1$ | $5 \cdot 10^{-3}$<br>$5 \cdot 10^{-2}$<br>$1 \cdot 10^1$ | $5 \cdot 10^{-3}$<br>$5 \cdot 10^{-2}$<br>$1 \cdot 10^1$ | $2.5 \cdot 10^{-3}$<br>$2.5 \cdot 10^{-2}$<br>5 | $3 \cdot 10^{-3}$<br>$3 \cdot 10^{-2}$<br>6 |
| **Veins conductance** $\gamma$<br>[s<sup>-1</sup>Pa<sup>-1</sup>] | $8 \cdot 10^{-6}$ | $8 \cdot 10^{-6}$ | $8 \cdot 10^{-6}$ | $8 \cdot 10^{-6}$ | $6 \cdot 10^{-6}$ | $6 \cdot 10^{-6}$ |
| **Right atrium pressure** $p_{ra}$<br>[mmHg] | 2 | 2 | 2 | 2 | 8 | 8 |
| **LAD coupling coefficient** $\alpha$<br>[m<sup>3</sup>s<sup>-1</sup>Pa<sup>-1</sup>] | $3 \cdot 10^{-9}$ | $3 \cdot 10^{-9}$ | $9 \cdot 10^{-9}$ | $6 \cdot 10^{-10}$ | $3 \cdot 10^{-10}$ | $9 \cdot 10^{-9}$ |
| **LCX coupling coefficient** $\alpha$<br>[m<sup>3</sup>s<sup>-1</sup>Pa<sup>-1</sup>] | $6 \cdot 10^{-10}$ | $1 \cdot 10^{-9}$ | $6 \cdot 10^{-10}$ | $1 \cdot 10^{-9}$ | $6 \cdot 10^{-10}$ | $6 \cdot 10^{-10}$ |
| **RCA coupling coefficient** $\alpha$<br>[m<sup>3</sup>s<sup>-1</sup>Pa<sup>-1</sup>] | $3 \cdot 10^{-10}$ | $3 \cdot 10^{-10}$ | $3 \cdot 10^{-10}$ | $3 \cdot 10^{-10}$ | $9 \cdot 10^{-9}$ | $9 \cdot 10^{-9}$ |
| **Calibration error** $E_{CAL}$ [%] | 1.3 | 2.4 | 0.8 | 1.5 | 8.1 | 1.8 |

### 3.2 Overview of digital twin results

Here we report the results of one representative patient (P4, belonging to *validation patients*) to highlight the effective potential of the digital twin in quantitatively describe the coronary and myocardial hemodynamics. To this purpose, we run the digital twin simulation with the parameters corresponding to the patient’s group A-II and post-processed as detailed in Section 2.5. In Fig. 6, we report a detailed analysis of the hemodynamics.

**Fig. 6:**
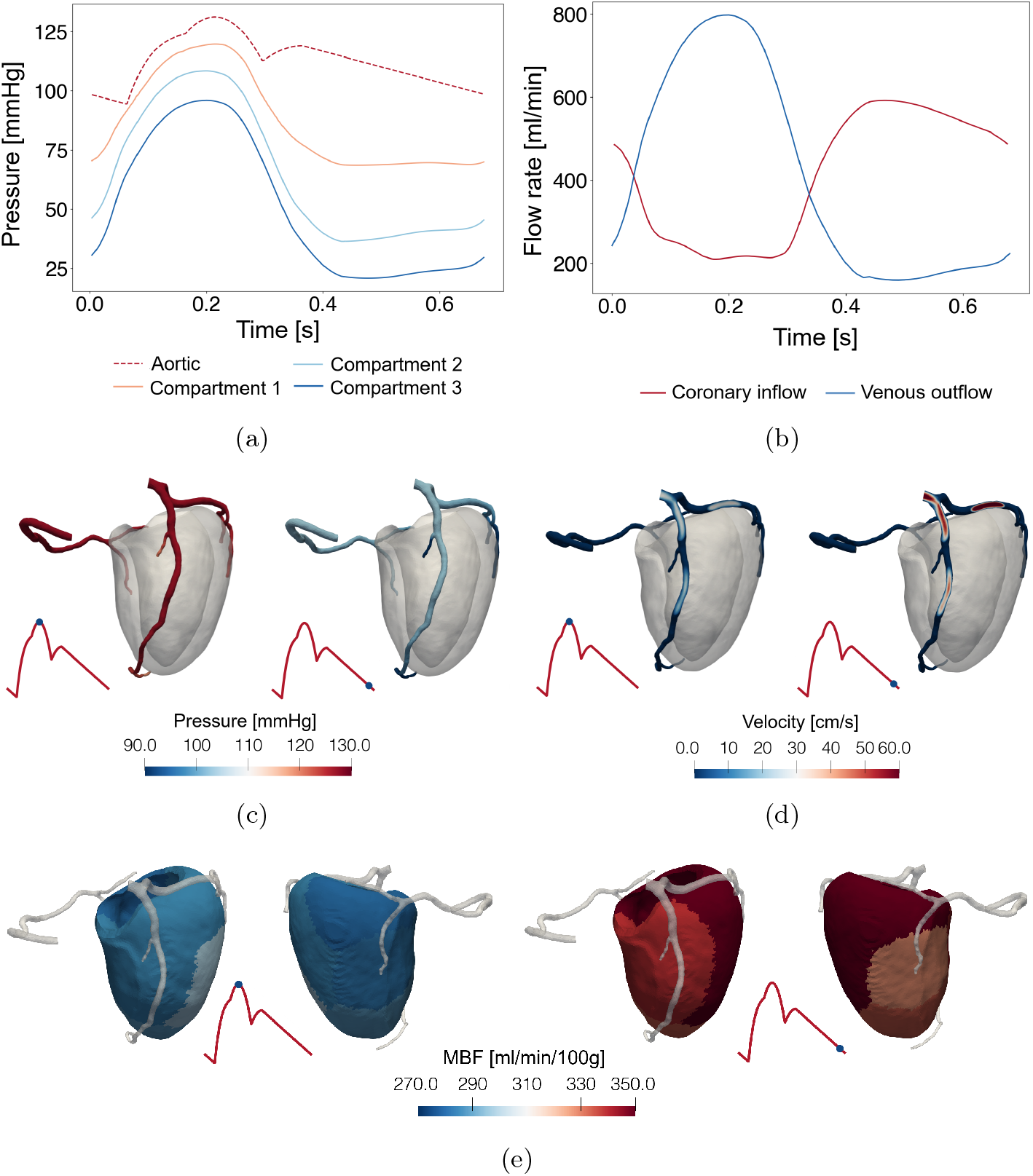
a) Prescribed aortic pressure and computed in-space-averaged pressure of the three myocardial compartments corresponding to small arteries, arterioles and capillaries, respectively; b) Arterial and venous coronary flow rate computed at the coronary inlet and at the third myocardial compartment, respectively; c) Coronary blood pressure at systole (left) and diastole (right); d) Coronary blood velocity at systole (left) and diastole (right); e) 3D MBF map averaged on each perfusion region at systole and diastole: for each instant the inferior view (left) and anterior view (right) are displayed. Computational results of P4’s digital twin, for figures c, d and e the time instant is indicated by a blue dot on the arterial pressure curve aside in red.

First, we plot the time evolution of the prescribed aortic pressure and of the computed in-space-averaged pressure in the three Darcy myocardial compartments (Fig. 6a). As expected, going from the first to the third compartment we observe that the pressure decreases and that its waveform tends to be more similar to the intramy-ocardial pressure (Fig. 4). The in-time-averaged pressure in the three compartments is 86.7, 64.8, 50.6 mmHg; this values are in good agreement with experimental measures of 88 and 68 mmHg reported in [38] for the first and second compartment, respectively.

From Fig. 6b, we can clearly see that the model is able to reproduce the physiological phasic flow of the coronary arteries, where we have high coronary inflow during diastole and a decrease of inflow during the systolic phase due to the myocardium contraction, while for the venous outflow we observe a standard systolic flow rate.

In Fig. 6c and Fig. 6d we report the pressure and velocity in the coronary arteries, both during systole (left) and diastole (right). At systole we have larger values of the pressure than at diastole, accompanied however by a lower velocity. This is due to the smaller pressure gradient between base and apex experienced at the systole with respect to the diastole (5.3 mmHg vs. 23.6 mmHg). The velocity values found are also compatible with experimental measures [39] where the average peak velocity is around 48 cm/s.

Finally, in Fig. 6e, we display the 3D map of the MBF averaged in space within each perfusion regions both at systole and diastole. As expected, the MBF is higher during diastole, when perfusion of the myocardium takes place, while during systole is lower due to myocardial contraction. Even tough the digital twin is capable of computing MBF over time, in the next section we will concentrate on the time-averaged MBF, as we need to perform a comparison with stress-CTP measures in order to validate the model (see Section 3.3).

### 3.3 MBF prediction of the validation patients

In this section we report the results of the digital twin simulations for the *validation patients* and we analyze the results with the aim of validating our model against clinical measures from stress-CTP.

In Fig. 7 we collected the stress-CTP measured maps (line M) and the digital twin MBF maps (line S) for the *validation patients*. Patients belonging to group B (severely stenotic) are characterized by generally lower MBF than patients from group A. In addition to the 3D MBF maps, in Table 5 we reported, for each patient, the validation error *E*_*V AL*_, defined in Section 2.5. The error featured by 20 patients over 28 is less than 10% (in green in Table 5), and in 14 of them the error is less than 5%. As expected, group A presents far better results than group B, due to the increased functional and geometrical complexity of the latter. In any case, we have good results also for group B, with 9 over 16 patients characterized by *E*_*V AL*_ *<* 10% and with an error always less than 33%. The average *E*_*V AL*_ among the patients is 2.6% for A-I, 6.2% for A-II, 4.3% for A-III, 4.8% for A-IV, 12.7% for B-I, and 9.9% for B-II.

**Table 5:** The value of *E*_*V AL*_ computed by (1) is reported for each patient. We highlight in green errors under 10%, in yellow errors between 10% and 15%, in orange errors between 15% and 20%, and in red errors over 20%.

| A-I |  | A-II |  | A-III |  | A-IV |  | B-I |  | B-II |  |
| --- | --- | --- | --- | --- | --- | --- | --- | --- | --- | --- | --- |
| P | $E_{VAL}[\%]$ | P | $E_{VAL}[\%]$ | P | $E_{VAL}[\%]$ | P | $E_{VAL}[\%]$ | P | $E_{VAL}[\%]$ | P | $E_{VAL}[\%]$ |
| P1 | 3.5 | P3 | 0.0 | P7 | 0.0 | P9 | 9.7 | P13 | 5.8 | P20 | 25.6 |
| P2 | 1.6 | P4 | 0.0 | P8 | 8.6 | P10 | 1.8 | P14 | 13.6 | P21 | 32.8 |
|  |  | P5 | 0.0 |  |  | P11 | 7.7 | P15 | 30.8 | P22 | 0.0 |
|  |  | P6 | 24.8 |  |  | P12 | 0.0 | P16 | 16.7 | P23 | 4.7 |
|  |  |  |  |  |  |  |  | P17 | 16.9 | P24 | 13.6 |
|  |  |  |  |  |  |  |  | P18 | 0.0 | P25 | 1.9 |
|  |  |  |  |  |  |  |  | P19 | 5.0 | P26 | 6.7 |
|  |  |  |  |  |  |  |  |  |  | P27 | 3.7 |
|  |  |  |  |  |  |  |  |  |  | P28 | 0.0 |

**Fig. 7:**
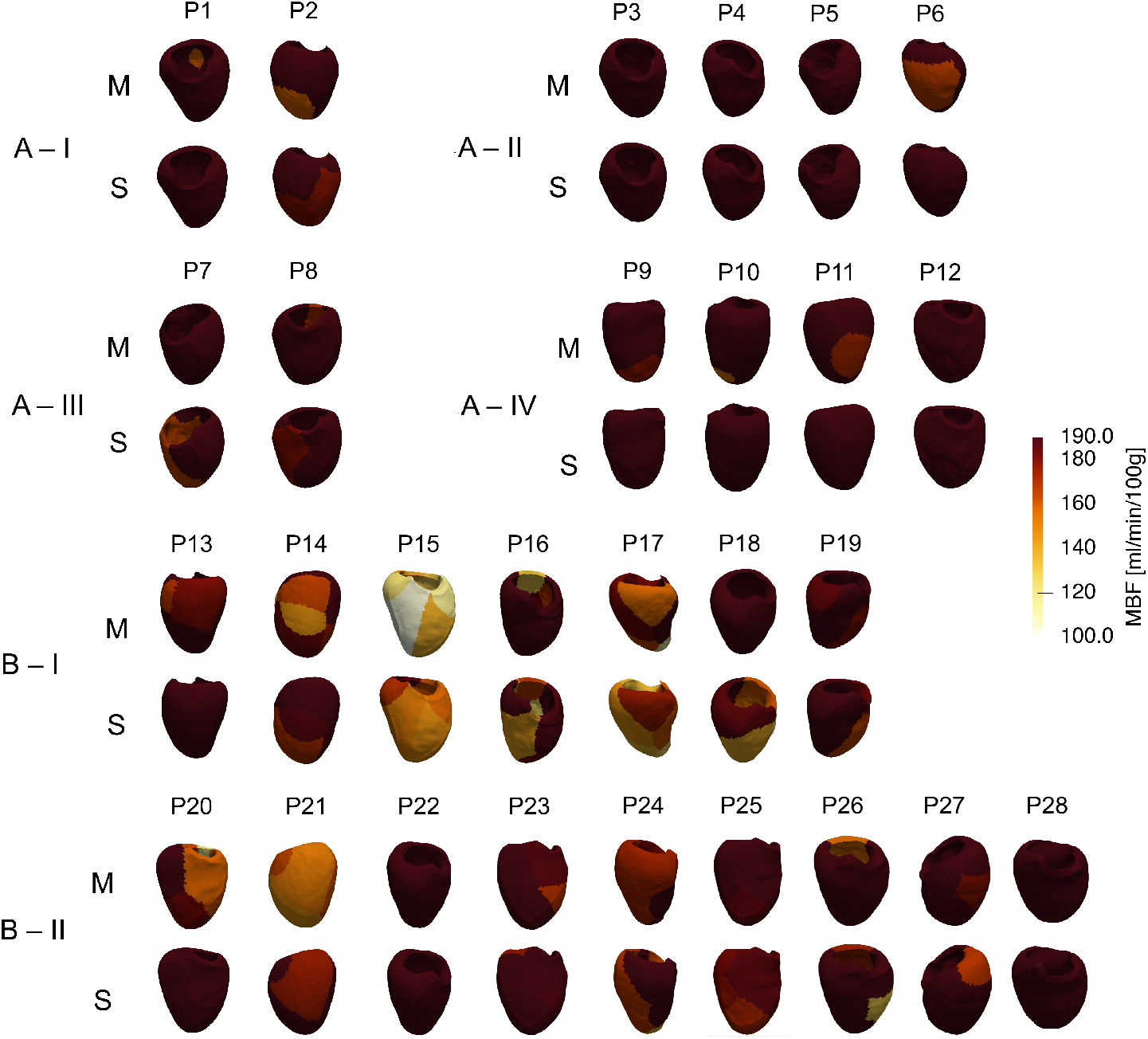
Stress-CTP measured MBF maps (line M) and digital twin simulated ones (line S) of the 28 validation patients. The range of the colorimetric scale highlights the regions used for *E*_*V AL*_ calculation.

To assess the accuracy of the model we perform a statistical analysis on the *validation patients* results. Specifically, we compare both the global in-space-averaged MBF (*M* and *S*) and the MBF averaged over the perfusion regions (*M*_*j*_ and *S*_*j*_). Specifically, in Fig. 8a we report *M* and *S* values (see Section 2.4) and we evaluate the corresponding correlation, obtaining a statistically significant (*p* = 0.001) moderate correlation (*R*^2^ = 0.58). Notice that the points are evenly distributed along the identity line (dashed line) with a neat separation between group A and B in terms of amount of perfusion, although we observe some dispersion. We did the same also for *M*_*j*_ and *S*_*j*_ for any perfusion region *j* (see Fig. 8b). Also in this case we find statistical significance (*p* = 5 *·*10^*−*14^) and moderate correlation between measure and the prediction (*R*^2^ = 0.49); additionally, we still observe an uniform distribution of the points along the identity line with a more evident dispersion for high values of *M*_*j*_ and *S*_*j*_.

**Fig. 8:**
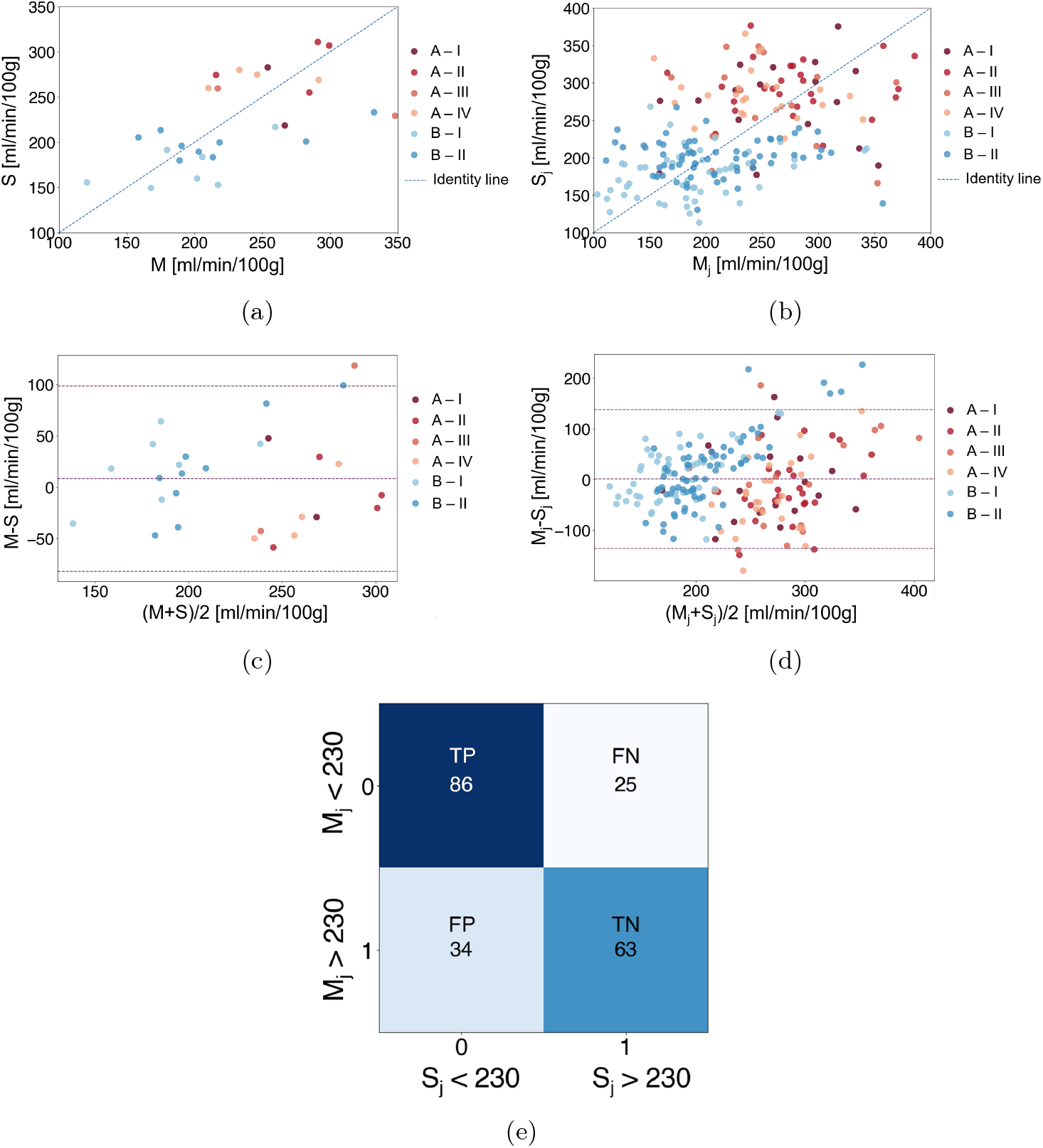
a) Correlation between *M* and *S*, each dot represents a patient; b) Correlation between *M*_*j*_ and *S*_*j*_, each dot represents a region; c) Bland-Altman analysis for *M* and *S*, each dot represents a patient (mean difference = 8.4, upper limit = 98.6, lower limit = −81.7); d) Bland-Altman analysis for *M*_*j*_ and *S*_*j*_, each dot represents a region (mean difference = 1.5, upper limit = 138.5, lower limit = −135.6); e) Confusion matrix obtained using as threshold 230 ml/min/100g for *M*_*j*_ and *S*_*j*_.

To further evaluate the agreement between the stress-CTP measures and the digital twin predictions we observe the results of the Bland-Altman tests, see Fig. 8c and Fig. 8d. In both cases the test delivers positive outcome, in particular we observe the absence of a systematic bias as the mean difference is close to 0 (8.4 ml/min/100g for *M − S*, 1.5 ml/min/100g for *M*_*j*_ *− S*_*j*_). The agreement limits define a range of the differences that is between −81.4 and 98.6 and between −135.6 and 138.5 for (M - S) and (M_*j*_ - S_*j*_), respectively, which we deem acceptable at this stage.

Finally, we want to evaluate the capability of the digital twin to predict a possible risk of perfusion impairment. To this aim we perform a classification test based on the confusion matrix analysis, and we establish that a region is at risk for perfusion impairment when its average MBF is below *η* = 230 ml/min/100g. In Fig. 8e we report the corresponding confusion matrix; we obtain a TPR = 0.77 and a TNR = 0.65. Moreover, precision = 0.72 and accuracy = 0.72. All in all, we observe that, despite the proposed digital twin is still being developed and this work constitutes the first attempt at his validation, the model performs moderately well.

## 4 Discussion

In this work, we defined our digital twin framework as the combined use of patient-specific reconstructed geometries, personalized boundary conditions, and a suitably calibrated computational model. The original outcome of the work is the prediction of myocardial blood flow for patient-specific cases and the application of our digital twin to a wide cohort of patients with the final aim of providing a first step towards its validation in view of clinical applications.

For the model calibration we divided the patients in six groups (see Section 3.1), according to the introduction of a cut-off (70%) for the stenosis degree. This choice lays in the association between ischemic lesions and a high degree of stenosis [27, 40]. Additionally, we considered also the volume ratios *R*_*L*_ and *R*_*L/R*_, since we hypothesized that the distribution of the myocardial perfusion regions over the ventricle volume has an impact on MBF prediction, as in our model their dimensions are linked to the radius and to the distribution of the main coronary arteries over the myocardium.

For each of the six groups, the calibration was performed by using one selected patient for each group, randomly chosen, and the validation was carried out by applying the calibrated model to the other patients (*validation patients*). From Table 5, we observe that the average MBF values obtained from the digital twin are in good agreement with the ones extracted from the stress-CTP maps, even if local discrepancies could be observed when comparing the measured MBF maps with the estimated ones, as exemplified in Fig. 7. However, our primary goal is to discriminate between lesions and healthy regions, so that an overly detailed analysis, based on a pointwise evaluation of MBF values, rather than on their significant ranges, could be here too constraining for our purpose.

The good agreement of the digital twin results with the stress-CTP measures is corroborated also by the statistical analysis; specifically the Bland-Altman analysis (Fig. 8c and Fig. 8d) suggests a good agreement of the results with the clinical measurements, especially because of the low bias (i.e. the average of the differences) observed. Also the confusion matrix analysis in the same figure provides positive outcomes, in particular the values of TPR, precision and accuracy are relevant (*>*0.7) highlighting that our model would be able to discriminate between at risk (positive) and non at risk (negative) patients. As for the lower TNR, this is due to more false positive than false negative cases in our classification, suggesting that tool works in a safety condition, i.e. it is more prone to suggest treating a patient that was not at risk rather than not treating an at-risk patient.

Observe that, the confusion matrix analysis is based on the chosen threshold (230 ml/min/100g). In the clinical practice, inducible ischemia is identified in dynamic stress-CTP from the presence of MBF values lower than [75 - 120] ml/min/100g in a region *≥* 25% of transmural myocardial thickness within a specific coronary territory [26, 41, 42]. However, in our case we did not focus on local estimation of MBF, rather only on average ones over the perfusion regions, where high variations of MBF could be experienced due to the presence of induced ischemia. Thus, we selected our threshold as follows: *η* = (75 + *MAX*)*/*2, where MAX (in our case 385 ml/min/100g) is a reference maximum MBF, obtained as the average among the patients of the maximum values. This choice guarantees that, if the estimated MBF is larger than *η*, supposing a linear variation of MBF between epicardium and endocardium, then each point of the region will have a MBF higher than 75 ml/min/100g.

The choice of estimating average MBF quantities for each perfusion region implies not having locally detailed information on perfusion, particularly in the direction from the epicardial to the endocardial surface. This reflects on the way we visualize results in Fig. 7; we chose a range characterized by a maximum that is the cut-off identified in the definition of *E*_*V AL*_ and a minimum that is a value above the clinical threshold (75 ml/min/100g). The latter, indeed, is never reached by average MBF quantities precisely because of the lack of locally detailed information on small specific regions in which this value can be observed.

Nevertheless, a pointwise analysis of MBF was out of the scope of this work, which aimed at providing a first validation of the digital twin on cardiac perfusion; so average quantities, estimated on well defined regions which cover the whole myocardium, are enough for our purposes. Moreover, our choice to work with average quantities allowed us to go towards the definition of indicators useful to the diagnostic application of our tool. For example, identifying each perfusion region with a single value of MBF enables to quickly recognize regions affected by a perfusion defect and, consequently, to identify the coronary artery possibly responsible for it.

Regarding the calculation of the error between the clinical and simulated MBF, we first observe that *E*_*V AL*_ is characterized by three types of approximations introduced by the stress-CTP machine (slice thickness), by the computational discretization (element length and time step) and by the method we used to post-process and compare the prediction to the stress-CTP measure. Regarding the latter, we project the stress-CTP map onto the segmented myocardium; however, since these do not always overlap perfectly, this juxtaposition produces an uncertainty in the error calculation, that is performed considering the projected map values.

All in all, the proposed digital twin is able to provide a calculation of 3D MBF maps in about three hours. We want to remark that this is, to our knowledge, the first work on a digital twin for MBF prediction to report such positive results with a blinded calibration (i.e. not influenced by clinical measures) and on a cohort of quite a few patients. From the 3D MBF maps we are able to extract both generic data (e.g. the global in-space-averaged MBF *S*) but also detailed information about the perfusion in each identified region (*S*_*j*_).

There are still some limitations that we list here:

- The lack of a right ventricle model, which could be used to better reproduce the flow repartition between right and left coronary arteries. This issue will deserve particular attention in future developments of the present work;
- We do not model the deformation of the left myocardium during contraction, we only reproduce the effect of systolic occlusion of intramyocardial coronaries by means of our surrogate model. Future work could account for this by introducing a poroelastic model or by prescribing displacement obtained with time resolved images (which however for stress-CTP are not available nowadays);
- We divide the myocardium in 7/8 perfusion regions, these could be not enough to represent the complex dynamics of myocardial perfusion. However, the use of such number of regions allows us to have a fast tool to be used in the clinical practice while increasing the number of regions would increment the computational time;
- Our perfusion model is able to identify and quantify perfusion defects due to coronary stenosis but not due to an impaired microcirculation. Possible ways to overcome this limitation will be to supply the model with additional parameters to describe this phenomenon;
- The calibration is carried out by selecting randomly six representative patients; the impact of this selection on the results remains to be evaluated. Moreover, the cutoff values of the ratios *R*_*L*_ and *R*_*L/R*_ are defined without performing a sensitivity analysis. Such analysis to evaluate the impact of these choices will be mandatory in future works;

We estimate average MBF quantities, so we are not able to give detailed information about local characteristics of perfusion, specifically in the transmural direction. This is surely an aspect to be improved in the future, even if, to the scope of this work, average quantities are considered suitable.

In conclusion we can state that our digital twin is proving to be a new effective diagnostic tool, especially for its capability of computing MBF average indicators that already show a good agreement with the clinically measured ones and that are able to sum up information about myocardial perfusion ready to be used by the clinicians. Finally, we want to stress out that this is the first time, to our knowledge, that a model for MBF prediction which features 3D modeling of both coronaries and myocardium hemodynamics has been used on such a sizable cohort.

## Data Availability

All data produced in the present study are available upon reasonable request to the authors

## Aknowledgements

This study was funded and E.C. was supported by the European Union– Next Generation EU – NRRP M6C2 – Investment 2.1 Enhancement and strengthening of biomedical research in the NHS-Project: PNRR-POC-2022-12376500, CONCERTO.

G.N was supported by the National Plan for NRRP Complementary Investments (PNC, established with the decree-law 6 May 2021, No. 59, converted by law No. 101 of 2021) in the call for the funding of research initiatives for technologies and innovative trajectories in the health and care sectors (Directorial Decree no. 931 of 06-06-2022) – project no. PNC0000003 – Advanced Technologies for Human-centred Medicine (ANTHEM). This work reflects only the authors’ views and opinions: neither the Ministry for University and Research nor the European Commission can be considered responsible for them.

G.M.P. and C.V. are members of the INdAM group GNCS “Gruppo Nazionale per il Calcolo Scientifico” (National Group for Scientific Computing).

C.V. has been partially supported by: i) the European Union-Next Generation EU, Mission 4, Component 1, CUP: D53D23018770001, under the research project MIUR PRIN22-PNRR n.P20223KSS2, ”Machine learning for fluid structure interaction in cardiovascular problems: efficient solutions, model reduction, inverse problems”, ii) the Italian Ministry of Health within the PNC PROGETTO HUB LIFE SCIENCE - DIAGNOSTICA AVANZATA (HLS-DA) ”INNOVA”, PNCE3-2022-23683266–CUP: D43C22004930001, within the ”Piano Nazionale Complementare Ecosistema Innovativo della Salute” - Codice univoco investimento: PNCE3-2022-23683266; iii) the Italian research project MIUR PRIN22 n.2022L3JC5T ”Predicting the outcome of endovascular repair for thoracic aortic aneurysms: analysis of fluid dynamic modeling in different anatomical settings and clinical validation”; iv) Italian Ministry of Health within the project “CAL.HUB.RIA” - CALABRIA HUB PER RICERCA INNOVATIVA ED AVANZATA. Code: T4-AN-09, CUP: F63C22000530001.

## Author contributions

Acquisition of the clinical data: A.B., G.P. Methodology: E.C., G.M.P. Conceptualization: C.V., G.P. Software: G.N., A.R. Formal analysis and investigation: E.C. Interpretation of the results: E.C. Writing—Original draft preparation: E.C., C.V. All authors reviewed the manuscript.

